# Immunogenicity and adverse events of priming with inactivated whole SARS-CoV-2 vaccine (CoronaVac) followed by boosting the ChAdOx1 nCoV-19 vaccine

**DOI:** 10.1101/2021.11.05.21264700

**Authors:** Surakameth Mahasirimongkol, Athiwat Khunphon, Oraya Kwangsukstid, Sompong Sapsutthipas, Minkwan Wichaidit, Archawin Rojanawiwat, Nuanjun Wichuckchinda, Wiroj Puangtubtim, Warangluk Pimpapai, Sakulrat Soonthorncharttrawat, Asawin Wanitchang, Anan Jongkaewwattana, Kanjana Srisutthisamphan, Daraka Phainupong, Penpitcha Thawong, Pundharika Piboonsiri, Warittha Sawaengdee, Thitiporn Somporn, Kanokphon Ritthitham, Supaporn Chumpol, Nadthanan Pinyosukhee, Rattanawadee Wichajarn, Panadda Dhepaksorn, Sopon Iamsiritahworn, Supaporn Phumiamorn

## Abstract

**Background:** Responding to SARS-CoV-2 Delta variants escaped the vaccine-induced immunity and waning immunity from the inactivated whole virus vaccine, Thailand recently proposed a heterologous inactivated whole virus vaccine (CoronaVac) viral vector vaccine (ChAdOx1 nCoV-19) prime-boost vaccine regimen(I/V). This study aims to evaluate the immunogenicity and adverse events of this regimen by comparison with homologous CoronaVac, ChAdOx1 nCoV-19, and convalescent serum.

**Method:** Immunogenicity was evaluated by the level of IgG antibodies against the receptor-binding domain of the SARS-CoV-2 spike protein (S1 subunit) (anti-S RBD). At 2 weeks following the second dosage, a selection of random samples was tested for plaque reduction neutralisation (PRNT) and Pseudotype-Based Microneutralization test (PVNT) against SARS-CoV-2 variants of concern (VOCs). The safety profile of heterologous CoronaVac-ChAdOx1 nCoV-19 prime-boost vaccine regimen was described by interviewing at the 1-month visit.

**Result:** Between April to August 2021,426 participants were included in the study, with 155 obtaining CoronaVac-ChAdOx1 nCoV-19(I/V),32 obtaining homologous CoronaVac(I/I),47 obtaining homologous ChAdOx1 nCoV-19(V/V),169 with history covid-19 infection. Geometric mean titers (GMTs) of anti-S RBD level in the I/V group compare 2wks and 4 wks (873.9 vs 639,p=0.00114).At 4 wks, GMTs of anti-S RBD level in I/V group was 639, 95% CI 63-726,and natural infection group 177.3, 95% CI 42-221, and V/V group 211.1, 95% CI 77-152, and I/I group 108.2, 95% CI 77-152; all p<0.001).At 2 wks, The GMTs of 50%PRNT of 19 sampling from the I/V group is 434.5, 95% CI 326-579, against wild type and 80.4, 95% CI 56-115, against alpha and 67.4, 95% CI 48-95, against delta and 19.8, 95% CI 14-30, against beta; all p<0.001. At 2 wks, The GMTs of 50%PVNT of 15 sampling from the I/V group is 597.8, 95% CI 368-970, against wild type and 163.9, 95% CI 89-301, against alpha and 157.7, 95% CI 66-378, against delta. The AEs in the I/V schedule were well tolerated and generally unremarkable.

**Conclusion:** The I/V vaccination is a mixed regimen that induced higher immunogenicity and shall be considered for responding to Delta Variants when only inactivated whole virus vaccine and viral vector vaccine was available.

## Background

In lower and middle-income countries, where the access to the COVID-19 vaccine is limited in 2021, the estimated total COVID19 vaccine in 2021 is 4 billion doses while the demand is around 8 billion doses. Lower and middle-income countries relied on the COVID-19 vaccine such as the inactivated whole virus vaccine, CoronaVac^1^ (Sinovac Biotech, Beijing, China), and viral vector vaccine such as ChAdOx1 nCOV-19 vaccine^2^.

The heterologous prime-boost vaccination has been reported to be induced higher immunogenicity for various vaccine^3^ and routinely practice in the annual vaccination program such as influenza, or in the vaccination program, where vaccination of different manufacturing platforms is considered replaceable. While only one covid vaccine was designed based on heterologous rAd26 and rAd5 vector-based COVID-19 vaccination, which enables a high level of immunogenicity^4^.Most of the vaccines were not utilized the heterologous vaccination concept because most manufacturers did not develop the vaccine from two different vaccine delivery platforms or manufacturing platforms. It is a generally higher cost for manufacturing vaccines with two manufacturing platforms. The outbreaks of Delta variants from the beginning of 2021 in India are devastating with estimates of 10 times higher number of deaths than the reported deaths due to its higher transmissibility, more severe and vaccine escape capability^5^. Based on these characteristics, the Delta variant was recognized as the variants of concerns (VOCs) by Public Health England, US Center for Disease Control, and World Health Organization (WHO). WHO issued global notices of Delta variant as the dominant variants and notification of the vaccination program to accelerate the implementation of any vaccine that is readily accessible. The outbreaks of the Delta variant in Indonesia occurred in the middle of 2021^6^. In July 2021, the Royal Society of Physicians in Indonesia reported the highest mortality rates from COVID19 in health care workers who were fully vaccinated with CoronaVac, this report^6^ prompted global concerns about the efficacy of inactivated viral vaccine against the Delta variants after several months post-vaccination.In some countries where the inactivated viral vaccine was used at the beginning of 2021, such as Bahrain, China, United Arab Emirates, and Egypt, the booster dose (third dose) by mRNA vaccine were offered to elderly and high-risk populations in the middle of 2021^7^ The immunogenicity study of inactivated whole virus vaccine in Thailand suggested that inactivated virus vaccine with Wuhan’s variant S protein provided low immunogenicity against the alpha and Delta variant ^8^. The Delta variant in Thailand was first detected in May 2021, from the construction workers site in the Laksi district of Bangkok, where workers from Myanmar and Cambodia worked with Thai nationals. Within 3 months duration, and multiple outbreaks of the Delta variant, it took over the Alpha variant which predominated in Bangkok from April to May, and became the dominant variants of COVID19 in Thailand in the first week of August 2021. Like other countries in Southeast Asia, Thailand began the COVID-19 vaccination program with CoronaVac and ChAdOx1 nCOV-19, viral vector vaccine from AstraZeneca, mainly due to the accessibility of the inactivated viral vaccine from China. While the vaccination program was implemented from March to May 2021, a group of individuals who experienced vaccine-induced adverse reactions from CoronaVac was offered the ChAdOx1 nCOV-19 as the only possible alternative to CoronaVac, and their immunogenicity was measured and reported^9^. Based on immunogenicity data in those who experienced AEs from CoronaVac, a heterologous vaccine schedule, priming with inactivated SARS-CoV-2 vaccine and followed with viral vector vaccine were proposed and adopted based on the neutralizing antibody data from this vaccine schedule by the national vaccination program on 12^th^ July 2021. Therefore, Thailand proposed the heterologous prime-boost regimen, a 21-day interval between the first dose inactivated whole virus vaccine (CoronaVac) and the second dose viral vector vaccine (ChAdOx1 nCoV-19 vaccine), from now we called this the “I/V” regimen, the only regimen that available as the preferred schedule along with the existing vaccine schedule for mitigating the mortality of COVID19 pandemic in Thailand. However, the immunogenicity in a larger number of samples and detailed information of common reactogenicity especially the adverse reaction of the second vaccination dose of ChAdOx1 nCoV-19 after inactivated viral vaccine had never been reported. Here we aim to determine the immunogenicity two weeks and four weeks after vaccination with the I/V schedule and describe the common adverse drug reactions from this I/V schedule.

## Method

### Study design

In this observational study, two groups of healthy individuals without a known history of covid-19 infection or exposure were recruited in this study. Their covid-19 infection or exposure were verified by interviewing and re-checking with the national registry of COVID-19. In I/V group was divided into two subgroups 1a and 1b. First, Group 1a, 30 participants, who received CoronaVac were those who experienced at least an adverse event following immunization and health care providers recommended that their second covid-19 vaccine should be switched to ChAdOx1 nCoV-19. Within the first half of 2021, only ChAdOx1 nCoV-19 is available in Thailand as the alternative to the inactivated viral vaccine. These patients were recruited from the Bangsue central vaccination center, the largest center for COVID-19 vaccination in Thailand. The group 1b of participants were enrolled from MOPH vaccination center vaccination site at the permanent secretary office of the Ministry of Public health, those who were scheduled to receive a second dose of inactivated viral vaccine were offered to switch their second dose vaccination to ChAdOx1 nCoV-19. The offer is based on the recommendation to I/V vaccination from the national vaccination academic subcommittee. The benefit of this I/V regimen was an opportunity to achieve higher immunogenicity than I/I regimens and in shorter duration intervals when compared to the V/V regimen, the risk was the possibility of a higher rate of adverse events than the I/I schedule.

The benefit and risks of second vaccination with ChAdOx1 nCoV-19 were explained to participants. The ethical approval of this study was approved by the ethical committee of the Department of Medical Sciences with approval number; MOPH 0625/EC060. Written informed consent was obtained from all participants.

Vaccination was performed according to the vaccination center guideline; the timing and lot of vaccines were recorded in the national vaccine information system called “Ministry of Public Health Vaccine Information Center (MOPH-IC)”.Immunogenicity analysis after the secondary vaccination at the Bangsue central vaccination center (BCVC) was offered as the test-based voluntarily. All participants provided written informed consent to have their immunogenicity and the adverse event included in this study.

All participants were invited to monitor immunogenicity at 2 weeks and 4 weeks after the second vaccination with ChAdOx1 nCoV-19.

The immunogenicity data from the fully vaccinated viral vector vaccine (V/V group) were retrieved from participants at BCVC.

The fully vaccinated inactivated virus vaccine (I/I group) was recruited from another vaccine study in DMSc.The convalescent serum from those with COVID-19 Delta variant infection from Laksi district was utilized for immunogenicity analysis on Natural infection in 2021 (NI group). Written informed consent was acquired from these two groups of fully vaccinated individuals and one group of natural infection 2021. The timing of samples collected from I/I groups and V/V groups was at 4 weeks after vaccination.

### Quantification of SARS-CoV-2 anti-S RBD antibody

The level of immunoglobulin class G (IgG) antibodies to the receptor-binding domain (RBD) of S1 subunit spike protein of SARS-CoV-2 was measured and quantified in human serum or plasma by using the ARCHITECT System (Abbott, Abbott Park, Illinois, USA) chemiluminescent microparticle immunoassay (CMIA) (SARS-CoV-2 IgG II Quant, Abbott Ireland, Sligo, Ireland) with measuring reportable range from 6.8 Abbott Arbitrary Unit (AU/mL) to 80,000.0 AU/mL (up to 40,000 AU/mL with onboard 1:2 dilution). Values higher than 50 AU/mL were considered positive. Based on the evaluated dilutions of the World Health Organization (WHO) International Standard (NIBSC Code 20-136) for anti-SARS-CoV-2 human immunoglobulin in WHO binding antibody unit (WHO BAU/mL) with the SARS-CoV-2 IgG II Quant assay with Abbott internal reference calibrators, the correlation between relationships of the AU/mL unit to the WHO (BAU/mL unit is at 0.142 × AU/mL) with the 0.999 correlation coefficient.

### Pseudotype-Based Microneutralization Assay against SARS-CoV-2

Pseudotype-Based Microneutralization Assay was carried out at the viral vaccine research center, National center for sciences and technology (NSTDA). Pseudotyped viruses (PVs) were produced in HEK293T/17 cells. HEK293T/17 producer cells were sub-cultured in 6-well plates (Thermo Fisher Scientific, Waltham, MA, USA) and co-transfected at 80-90% confluence with the p8.91 lentiviral packaging plasmid (500 ng), the pCSFLW firefly luciferase reporter plasmid (1 µg), and the pCAGGS plasmid encoding codon-optimized SARS-CoV-2 spike (1.5µg)^10^. The transfection was performed using polyethylenimine (PEI). The cell supernatants were collected 72 h before storage in microcentrifuge tubes at −80 °C.

Pseudotype-based microneutralization (pMN) assays were performed as previously described^10^ with some modifications. Briefly, human sera from SARS-CoV-2 seropositive and seronegative were subjected to two-fold dilution (starting from 1:40) in a white flat-bottom 96-well tissue culture-treated plate, and 50 µL of PV containing supernatant diluted in complete DMEM to give 5 × 10^5^ RLU equivalent was added per well. The plate was then centrifuged at 300× g for 3 min before incubation in a humidified cell-culture incubator for 1 h at 37 °C, 5% CO_2_. Subsequently, HEK293T cells stably expressing human ACE2 and TMPRSS2 (1.5 ×10^4^ cells/well) were mixed with the PV-serum complex before incubation in a humidified cell-culture incubator for 48 h at 37 °C in 5% CO_2_.

To measure luminescence activity, equal volumes of Bright-Glo™ reagent (Promega, Madison, WI, USA) and Phosphate Buffered Saline (PBS) were mixed and 25 µL added to each well; luminescence output was measured using a Synergy™ HTX Multi-Mode Microplate Reader (BioTek, Winooski, VT, USA) after 5 min incubation at room temperature. Analysis was performed by non-linear regression after normalization to 100% and 0% neutralization using GraphPad Prism Software.

### Plaque Reduction Neutralization Test (PRNT)

PRNT in this study is developed and tested by the Institute of Biological Products, WHO-contracted laboratory at the Department of Medical Sciences. Vero cells were seeded at 2x 10^5^ cells/well/ 3 ml and placed in 37°C, 5% CO2 incubator for 1 day. Test serum was initially diluted at 1:10, 1:40, 1:160, and 1:640, respectively. The SARS-CoV-2 virus was diluted in a culture medium to yield 40-120 plaques/well in the virus control wells. Cell control wells, convalescent patient serum, and normal human serum were also included as assay controls. The neutralization was performed by mixing the equal volume of diluted serum and the optimal plaque numbers of SARS CoV-2 virus at 37°C in the water bath for 1 hr. After removing the culture medium from Vero cell culture plates, 200 ul of the virus-serum antibody mixture were inoculated into monolayer cells and then rocked the culture plates every 15 min for 1 h. Three ml of overlay semisolid medium (containing 1% of carboxymethylcellulose, Sigma Aldrich, USA, with 1% of 10,000 units/ml Penicillin-10,000 ug/ml Streptomycin (Sigma, USA) and 10% FBS) were replaced after removing excessive viruses. All plates were incubated at 37OC, 5% CO2 for 7 days. Cells were fixed with 10% (v/v) formaldehyde then stained with 0.5% crystal violet in PBS. The number of plaques formed was counted in triplicate wells and the percentage of plaque reduction at 50% (PRNT50) was calculated. The PRNT50 titer of the test sample is defined as the reciprocal of the highest test serum dilution for which the virus infectivity is reduced by 50% when compared with the average plaque counts of the virus control and was calculated by using a four-point linear regression method. Plaque counts for all serial dilutions of serum were scored to ensure that there was a dose-response.

### Monitoring of adverse events after vaccination

Adverse events (AEs) were determined according to the SOP 43-05-17-CL-006 Handling and reporting of Adverse Events protocol, AEs were retrieved from questionnaires and/or telephone-based interviews. The registered nurse contacted all subjects, and none of them reported any serious adverse effects. The rates of each adverse drug reaction were reported. The Common Terminology Criteria for Adverse Events - Version 5.0 used to evaluate the severity of AEs.

### Statistical analysis

All the statistical analyses were performed using R (version 4.0.2) ^11^ and Rstudio (version 1.3.1093)^12^. The anti-SARS-CoV-2 RBD IgG level, PRNT_50_ titer, and PVNT_50_ titer were presented as geometric mean titers (GMTs) with 95 percent confidence intervals (CI).The difference of the antibody levels between vaccine groups: NI, I/I, I/V, V/V, at 4 weeks with between 2weeks and 4 weeks after having the I/V schedule were tested using the non-parametric (The Mann– Whitney U test)^13^ (available within the R package rstatix). The Wilcoxon signed-rank test, a non-parametric statistical hypothesis test for paired data ^13^, was applied to compare the PRNT_50_ titers and PVNT_50_ titers among selected SARS-CoV-2 variants. The Spearman’s rank correlation coefficient ^11^ (R function cor. test) was used to assess how well the relationship between SARS-CoV-2 variants and neutralization Assays: PVNT_50_ and PRNT_50_.

## Results

Between April to August 2021,403 participants were included in the study, with 155 obtaining CoronaVac-ChAdOx1 nCoV-19 (I/V group),32 obtaining homologous CoronaVac (I/I group),47 obtaining homologous ChAdOx1 nCoV-19(V/V group),169 with history covid-19 infection (NI group) (figure 1,2).In I/V group,155 participants were 76(49.3%) male and 79 (50.97%) female. The median age was 40 years (SD 9.4), 124 (80%) participants in 18–49 years and 31(20%) of participants are in 50-70 years.

The Demographic data between vaccination groups were described in Table 1.

### Immunogenicity profiles of I/V schedule compared with other vaccine schedules in Thailand

At 2wks visits, 149 participants were in the I/V group;6 participants missed their 2 wks visit but returned for visit at 4 weeks; geometric mean titers (GMTs) of anti-S RBD levels were at 873.9 BAU/mL (95% CI 768.4-993.8).

At 4 wks visit, 137 participants in I/V group, excluded 18 participants; 1 participant excluded due to HIV and low CD4 count, 2 participants infected with covid 19 after 2 weeks visit and 15 participants were lost to follow up; GMTs of anti-S RBD levels were at 639 BAU/ml (95% CI 63-726).

At 4 wks after the 3-6 weeks (mean 3.5 wks) interval between the two-dose CoronaVac, 32 participants with GMTs of anti-S RBD levels were 108.2 BAU/mL(95% CI 77-152).

At 4 wks after the 8-12 weeks interval between the two-dose ChAdOx1 nCoV-19, 47 participants with GMTs of anti-S RBD levels were at 211.1 BAU/mL (95% CI 162-249).

For the natural infection group, 169 participants with documented covid 19 infection from Laksi district with GMTs of anti-S RBD levels were 177.3 BAU/ml (95% CI 42-221). Most of the natural infection groups are asymptomatic individuals.

At 4 wks following the I/V schedule provided significantly higher GMTs of anti-S RBD level when compared with I/I, V/V schedule, and natural infection group(p<0.001).

### Antibody levels at 2 weeks after second vaccination compared with 4 weeks

-The antibody levels of the I/V schedule at 2 wks are significantly (p=0.00114) (873.9 BAU/mL, 95% CI 768.4-993.8) higher than the I/V schedule at 4 wks (639 BAU/mL, 95% CI 63-726). (Figure 4)

-The antibody levels of I/V schedule at 4 wks after second vaccination is significantly (p<0.001) (639 BAU/mL, 95% CI 63-726) higher than the V/V group (211.1 BAU/mL, 95% CI 162-249), NI group (177.3 BAU/mL, 95% CI 42-221), I/I group (108.2 BAU/mL, 95% CI 77-152). (Figure 3)

An exceptional participant with undetectable antibody was characterized for the low level of the Binding Antibody Unit at 2.4 AU/mL, due to high-risk occupation from the interview, the participant was subsequently counseled for the HIV screening and tested positive with an anti-HIV rapid screening test (SD BIOINE HIV1/2 3.0, Standard Diagnostics, Kyonggi-Do, South Korea) and the CD4 level, at 6.8 %. This patient was then referred to confirmatory diagnosis and treatment of HIV. This participant was excluded from further analysis.

### COVID19 breakthrough infection following immunization

During a telephone interview, two participants reported to the investigator that they couldn’t attend the 4 wks visit for blood samples collection due to their COVID19 situation that they experienced during the third week after the I/V schedule. These two participants in the age range 30-35 are living in the same household. BAU data at 2 weeks were as followed: the first participant had 380.3 BAU/ml whereas the second participant had 1,145.7 BAU/ml. The two participants were confirmed with COVID19 infection by RT-PCR at a private hospital, the first participant was diagnosed with COVID19 3 days earlier than the second participant and both of them experienced mild symptomatic covid 19 (with fever, malaise, and headache).

Their physician treated them with favipiravir, an antiviral agent, under a home isolation scheme, followed up chest radiography was unremarkable and reported no further complication.

### Neutralization antibody to variants of concern

The PRNT_50_ of 19 samples from the I/V schedule was analyzed and reported in figure 5 and table 7. While all 19samples demonstrated the neutralizing activity against the Alpha and Delta variant at > 10 IC cut-off, the neutralizing activity to Beta variants was achieved in 63% of these samples. In 19 participants from the I/V group were evaluated for their neutralizing antibody (Nab) by PRNT_50_ for 4 variants of SARS-CoV-2 (Wuhan-strain or wild type, Alpha variant, Delta variant, Beta variant). At 2 wks,the PRNT_50_ was the highest against wild type (434.5 BAU/mL, 95% CI 326-579),that significantly higher than to Alpha variant (80.4 BAU/mL, 95%CI 56-115),Delta variant (67.4 BAU/mL, 95% CI 48-95) and Beta variant(19.8 BAU/mL, 95%CI 14-30),respectively (p<0.001).

Moreover, their PRNT_50_ had the correlation coefficient with Ab to anti-s RBD level with wild type (ρ =0.82, p<0.001), alpha variant (ρ =0.66, p=0.0027), and Beta variant (ρ =0.074, p=0.76), delta variant (ρ =0.17, p=0.48) (Figure 7).

In addition, all participants (n=19/19) had PRNT_50_ titers ≥10 (Nab positivity cut-off) ≥10 (Nab positivity cut-off) against wild type, Alpha, and Delta strains. Participants (n=12/19,63%) had PRNT_50_ titers ≥10 against beta variant (Table 9).

### Pseudo Neutralization antibody to variants of concern (PVNT)

The PVNT_50_ of 15 samples from the I/V schedule was analyzed and reported in figure 6 and table 8. From the I/V group, 15 participants were evaluated for their neutralizing antibody (Nab) by PVNT_50_ for 3 variants of SARS-CoV-2 (wild type, Alpha variant, Delta variant). Beta variant for PVNT_50_ was not available at the time of this study. At 2 wks, the GMTs of PVNT_50_ is the highest against wild type (597.8, 95% CI 368-970) that significantly (p<0.001) greater than Alpha variant (163.9, 95% CI 89-301) and delta variant (157.7, 95% CI 66-378) (p<0.001). Furthermore, their PVNT_50_ were not significantly correlated with anti-s RBD level (ρ = 0.39, p=0.16), alpha variant (ρ =0.47, p=0.076) and delta variant (ρ =0.26, p=0.35) (Figure 8).

When considering the PVNT_50 titers_ ≥40 (Nab positivity cut-off), we demonstrated that all participants (n=15/15) achieved PVNT_50_ titers > 40 against wild type and Alpha variant. Participants (n=13/15,86.67%) had PVNT_50_ titer**s** ≥10 against beta variant (Table 9).

Additionally, the correlation between PVNT_50_ titer with PRNT_50_ titer were wild type (ρ =0.62, p=0.015), alpha variant (ρ =0.82, p=0.00022), and Delta variant (ρ =0.81, p=0.00044) (Figure 9).

### Adverse events following immunization

#### Reactogenicity to first dose CoronaVac

The reactogenicity population consisted of 155 individuals in the I/V group who got the second dosage of ChAdOx1 nCoV-19

The adverse events (AEs) to the first vaccine dose with CoronaVac in this study were described separately into two groups of samples. Group 1a was 30 participants from Bangsue Vaccination Center (BVC), who reported a higher rate of AEs from CoronaVac. Most AEs were mild and moderate symptoms such as immunization stress-related response (ISRR) (n=5, 16.67%), injected site pain (n=3, 10%), maculopapular rash (n=2, 6.67%), and others were described in Supplementary Table 2. Only 7 participants had severe AE included anaphylaxis (n=5, 16.67%), stroke (n=1, 3.33%) and central venous sinus thrombosis (n=1, 3.33%). Consequently, all participants in group 1a alter to receive the viral vector in the second dose due to the AE from the first vaccination (Table 2).

The group 1b were 125 participants from the MOPH vaccination center who received the I/V regimen based on the recommended vaccine schedule. This group has lower rates of reactogenicity from CoronaVac, the most common systemic reactions were feeling feverish (n=8, 6.4%), drowsiness (n=5, 4%), headache (n=2, 1.6%), and local AE such as injection site pain (n=1, 0.8%) (Table3).

### Reactogenicity to second dose ChAdOx1 nCoV-19

After ChAdOx1 nCoV-19 as second vaccine, in total 155 participants, the most common systemic AEs were feeling feverish (n=104,67.10%), headache (n=51,32.9%), myalgia (n=40,25.81%), and malaise and drowsiness (n=30,19.35%). The most common local responses such as injection site pain (n=54,35.06%) and other were described in Supplementary(Table4).

## Discussion

The higher level of antibody response and neutralizing activity at 4 weeks against the Delta variant following the I/V schedule is encouraging for the national vaccine program implementation.

In our study, heterologous I/V vaccination with a mean 3.5-week interval presents antibody levels at 4 wks that are significantly higher than V/V, I/I, and natural infection 2021. This result could be explained from additional support from study^14^, the author demonstrated in mice that the first dose of inactivated viral vaccine followed by adenovirus vectors vaccine induced a significantly prominent T-cell response than two doses of inactivated vaccines. These might help B cell greater differentiation into plasma B cell that could provide higher antibody response to specific antigen more two doses of inactivated vaccines^15^.

In addition, antibody levels of the I/I schedule at 4 wks were lower than natural infection 2021.This agreement with a prior study that S1-RBD-binding IgG titer of natural infection 2021 was higher than homologous CoronaVac vaccination^8^.

Additionally, the I/V schedule provided a higher Nab level at 2wks against Alpha variants than the Beta variant. This is compatible with a previous study utilizing a pseudovirus neutralization assay, serum from CoronaVac vaccinees collected 14 days following the second dose of the vaccine presented higher Beta resistance to neutralization than Alpha^16^. Moreover, this Nab level at 2wks against all VOCs decreased significantly compared to WT correlate with study^8^.

The Nab is highest to wild type variant, then to Alpha variant followed by delta variant, and the least Nab to Beta variant. This is consistent with a study that analyzed neutralization capacities by protein-based 206 ACE2 RBD competition assay was highest in wild-type, followed by alpha, delta, and Beta, respectively^17^.

Besides, participants in the I/V schedule resulted in high PRNT_50_ and PVNT_50_ titers against to delta variant. Immune protection against SARS-CoV-2 infection symptoms are highly correlated with levels of neutralising antibodies^18^.The AstraZeneca vaccines were shown to be 60% effective against the Delta variant after two doses^19^.

This evidence provides supporting information for this I/V vaccination schedule in Thailand. Availability of immunogenicity data from the I/V vaccination schedule is vital for consideration of the national vaccination program to consider the best vaccine schedule that could stimulate the higher level of immunogenicity. Since the immunogenicity study may not be feasible for every country, lower- and middle-income countries where inactivated viral vaccine and viral vector vaccine is available may use this evidence to support their decision to use alternative vaccination schedules. While multiple inactivated viral vaccines and viral vector vaccines are available, not all combinations of vaccine schedules produce satisfactory immunogenicity response, the early analysis in Thailand suggested that viral vector followed by inactivated viral vaccine-induced unsatisfactory immunogenicity^20^. While the correlation of protection for COVID19 vaccine has not been officially declared by World Health Organization, neutralizing activity to SARS-CoV-2 variants is likely related to the effectiveness of vaccination schedule ^21^. Each country can measure the immunogenicity of these vaccination schedules in the target population and utilized this information to guide the vaccination schedules at the national level. In countries with CoronaVac and ChAdOx1 nCoV19 are available like Thailand, using this heterologous vaccination schedule is likely to enable the highest level of immunogenicity to Delta variants from these two vaccines.

An individual with a low level of immune response after COVID vaccination with this I/V schedule was diagnosed with HIV infection with a low level of CD4. Two sources of information are used to make a prediction and identify the possible problem. Infection with HIV, which causes immunological damage to B-cell compartments and antibody production, might be substantially reduced humoral response to antigens and insufficient response of persons living with HIV to alternative vaccination, especially in those with low CD4 T-cell counts22.While immunogenicity is the proxy to the higher effectiveness, it is prudent to evaluate the safety and effectiveness of novel vaccine schedules after their introduction21. The introduction of the heterologous schedule of ChAdOx1 nCoV-19 followed by mRNA vaccination was evaluated for this vaccination schedule effectiveness (VE) in 100,000 fully vaccinated individuals, the real world VE of this vaccination schedule was at 88% (95% CI,83-92)^23^. The large-scale evaluation of safety and effectiveness for the I/V schedule is still carried out in Thailand, the results of VE study should come out in the next month.

The adverse reactions to vaccination in this study were described separately from two groups(1a,1b) of samples. Group 1a reported a higher rate of AEs from the first vaccination with the inactivated viral vaccine because this group received the viral vector due to the AEs from the first vaccination, the AEs rates from this group are not representative of AEs from the general population.Group 2b was retrieved from the general population that went to the vaccination center, the AEs from group 1b can be used as representative of general populations. The common AEs for the first vaccination with an inactivated viral vector was feeling feverish (n =8,6.4 %). According to our initial findings, the AEs after the second dose of ChAdOx1 nCoV-19 in the I/V schedule were well tolerated and generally safe. The adverse event profile reported here is comparable to previously reported adverse events following vaccination with ChAdOx1 nCoV-19^24^. Most participants had mild or moderate adverse effects and no report of serious AEs.The severity of local and systemic AEs of I/V schedule after the second dose of ChAdOx1 nCoV-19 in 7 days was mild to moderate corresponded to the reported AEs in phase II and phase III study of ChAdOx1 nCoV-19 (UK, Brazil, South Africa)^25^. The rate of systemic AEs, such as fever, recorded after the second dosage of ChAdOx1 nCoV-19 was higher (67.1%) than the earlier study (51%)^26^. However, Both systemic AEs (such as headache, malaise, myalgia) and local AE (such as injection site pain) were lower rates than this study^26^. This higher level of immunogenicity and the acceptable level of adverse reactions profile of this I/V vaccination schedule, this vaccination schedule is likely a better solution for countries that had access to both Inactivated whole virus vaccine and viral vector vaccine-like Thailand. The questions that remained to be answered are the duration of immunogenicity which may need following up study at 3 months, 6 months, 9 months, and 1 year. The decreasing levels of antibody level at 2 weeks to 4 weeks suggested that immunogenicity level may last between 3 to 6 months and the temporal relationship of effectiveness should be monitored carefully by the national vaccination program. The third dose is likely necessary for boosting immunogenicity after the I/V vaccine schedule and the types and timing of vaccine should be determined as the follow-up study from this cohort of vaccine recipients for providing a further recommendation.

There are several limitations to our research. First, our study did not collect comparison data on immunity levels at baseline before the study and after the first vaccine dose, hence, we could not exclude the effect of the previous infection on immunogenicity. However, the COVID-19 infection rate in Thailand was low before July 2021, there is a small possibility to include previously infected COVID-19 cases but these individuals should not affect the immunogenicity results in this study due to the immunity induced by infection were much higher in previously infected individuals compared with single-dose vaccination. A larger sample size is needed to detect uncommon adverse events such as vaccine-induced thrombotic thrombocytopenia and further studies should be done at the time of implementation of this vaccine schedule in the national vaccine program. Moreover, the PRNT_50_ and PVNT_50_ were assessed only in I/V regimen, limiting the comparability of the I/V regimen with other vaccine schedules, also the limited time for this study resulted in a smaller sample size that had neutralizing activities, however, the high level of neutralizing antibody suggested that the number of sample size is sufficient for the conclusion that I/V schedule is better than I/I schedule and comparable to V/V schedule.

Finally, our research offers crucial real-world proof of the safety and immunogenicity of heterologous CoronaVac-ChAdOx1 nCoV-19 immunization. The I/V vaccination is a mixed regimen that induced higher immunogenicity with a shorter duration to peak immunogenicity compared to the I/I schedule. In a situation where the viral vector vaccine is inadequate, we should consider this vaccine schedule for responding to Delta Variants. We propose that this initial assessment encourage future research of heterologous prime-boost vaccination regimens for COVID-19.

## Data Availability

All data produced in the present work are contained in the manuscript .
Surakameth Mahasirimongkol, MD, Ph.D.
Medical Life Sciences Institute, Department of Medical Sciences, Ministry of Public Health
88/7 Tivanond Road, Nonthaburi 11000, Thailand
Tel: (+66) 2951000, 0-2589-9580-8; Fax: (+66) 2-591-1707
. or

## Ethics Statement

The ethical approval of this study was approved by the ethical committee of the Department of Medical Sciences with approval number; MOPH 0625/EC060

## Conflicts of interest

All authors declare no conflict of interest.

## Funding

Department of Medical Sciences provided funding for this project. Studies, data collection, analysis, and interpretation, as well as manuscript preparation and the determination to submit the paper for publication, were all independent of funding sources.

## Acknowledgments

We would like to express our gratitude to the whole COVID-19 research team including all of the study participants. We thank Dr. Mark Simmerman for his critical data and recommendations.

## Notes

### Competing Interest Statement

The authors have declared no competing interest.

### Author Declarations

Research Ethics Committee,Department of Medical Sciences,Ministry of Public Health,Thailand has approved the following study which is to be carried out in compliance with ICH-GCP and 45 CFR 46.101(b)

### Summary of Updates

The names of two authors were repeated presented in the author list, the repeated names were removed.

