## Supplementary figure 1-9 for "Immunogenicity and adverse events of priming with inactivated whole SARS-CoV-2 vaccine (CoronaVac) followed by boosting the ChAdOx1 nCoV-19 vaccine"

### Table of figure

**Figure 1 : Enrollment diagram 1** 3

**Figure 2 : Enrollment diagram 2** 4

**Figure 3 :  Violin plots of level IgG anti-S RBD titer  in 4 group, Boxplots show geomean and IQRs.** 5

**Figure 4 : Violin plots  compare of level IgG anti-S RBD titer at 2 wks and 4 wks. Boxplots show geomean and IQRs.** 6

**Figure 5 :  Boxplots indicate geomean  $PRNT_{50}$ titer to virus variants and IQRs.** 7

**Figure 6 :  Boxplots indicate geomean  $PVNT_{50}$ titer to virus  variants and IQRs.** 8

**Figure 7 : Correlation between $PRNT_{50}$ titer with Ab to anti-s RBD level against  virus variants group** 9

**Figure 8 : Correlation between $PVNT_{50}$ titer with Ab to anti-s RBD level against  virus variants group** 10

**Figure 9 : Correlation between $PVNT_{50}$ titer with $PRNT_{50}$ titer against  virus variants group** 11

**Figure 1 : 155 participant were divided in 2 group for immunogenicity and adverse events analysis at 2 weeks and 4 weeks after second dose vaccination**

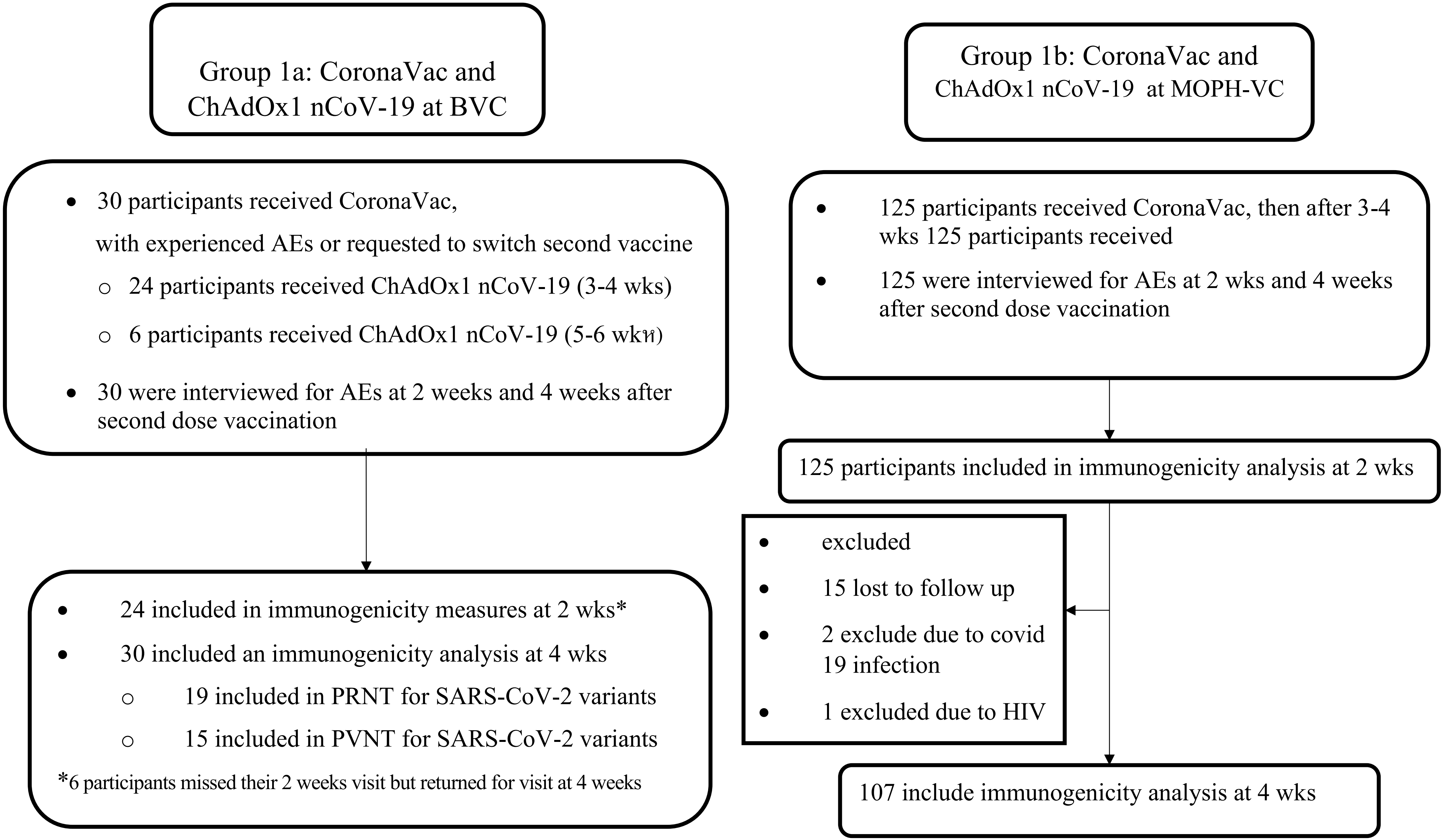

**Figure 2 : 248 participant from 3 group ( CoronaVac/ CoronaVac, ChAdOx1 nCoV-19/ ChAdOx1 nCoV-19, natural infection 2021) for immunogenicity at 4 weeks after second dose vaccination**

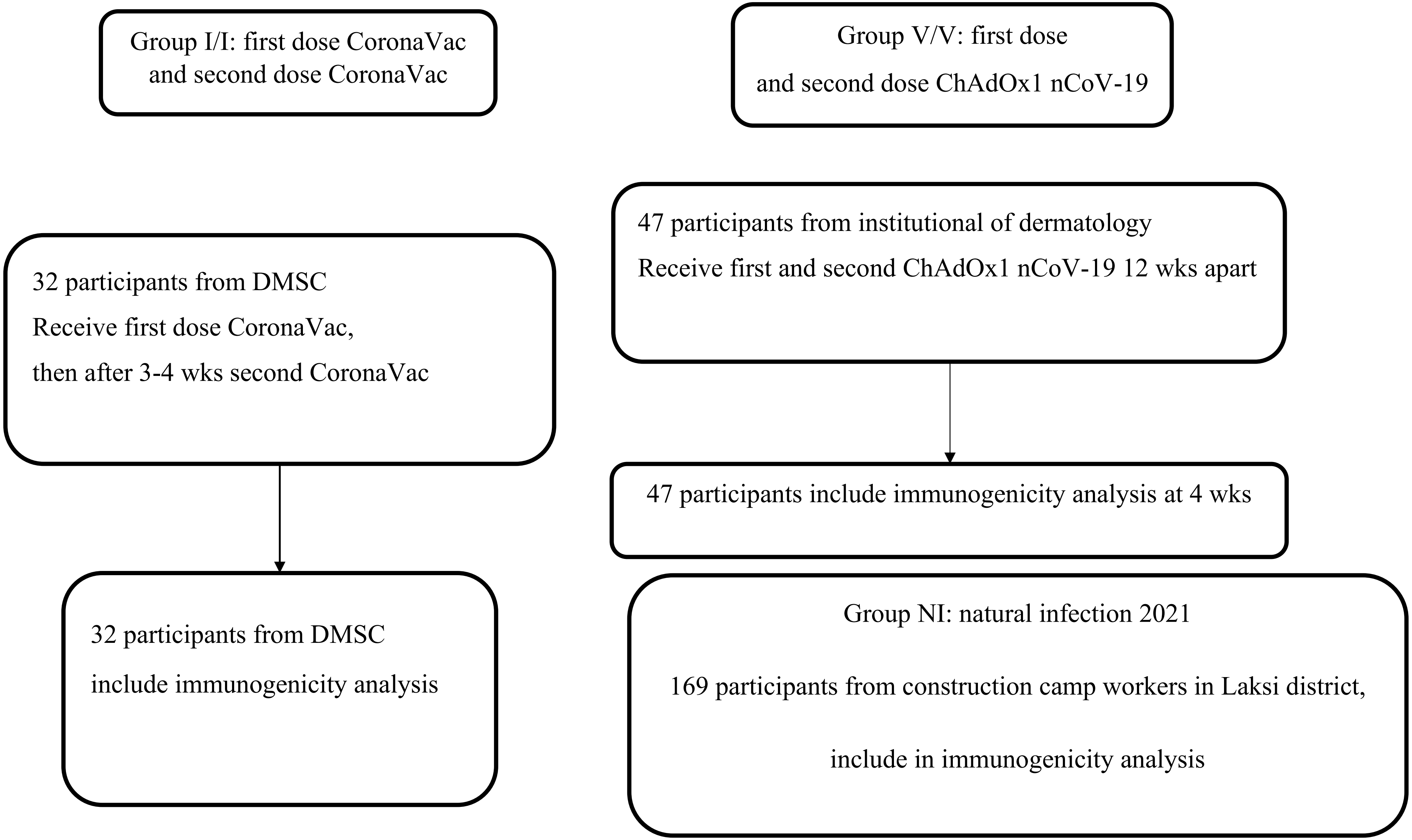

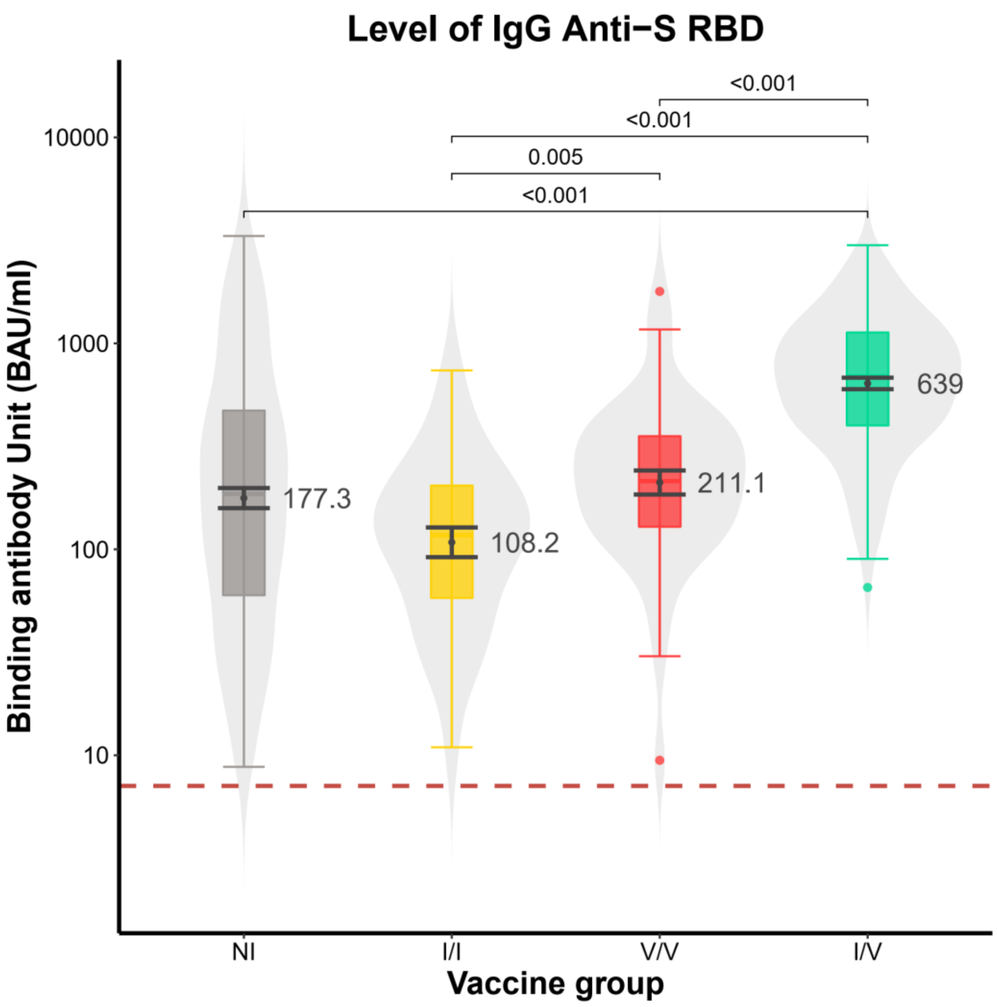


**Figure 3: Violin plots of level IgG anti-S RBD titer in 4 group (NI=natural infection 2021, I/I=Coronavac/ Coronavac, V/V=ChAdOx1 nCoV-19/ ChAdOx1 nCoV-19, I/V= Coronavac/ ChAdOx1 nCoV-19).Boxplots show geomean and IQRs. serum collection at 4 wk in 3 group(I/I,V/V,I/V).I/V group was significantly (p<0.0001) ( 639 BAU/mL, 95% CI 63-726) higher than V/V (211.1 BAU/mL, 95% CI 162-249), NI group( 177.3 BAU/mL, 95% CI 42-221) (p<0.0001) ,I/V ( 108.2 BAU/mL, 95% CI 77-152 ) (p<0.0001) by using the non-parametric the Mann–Whitney U test.**

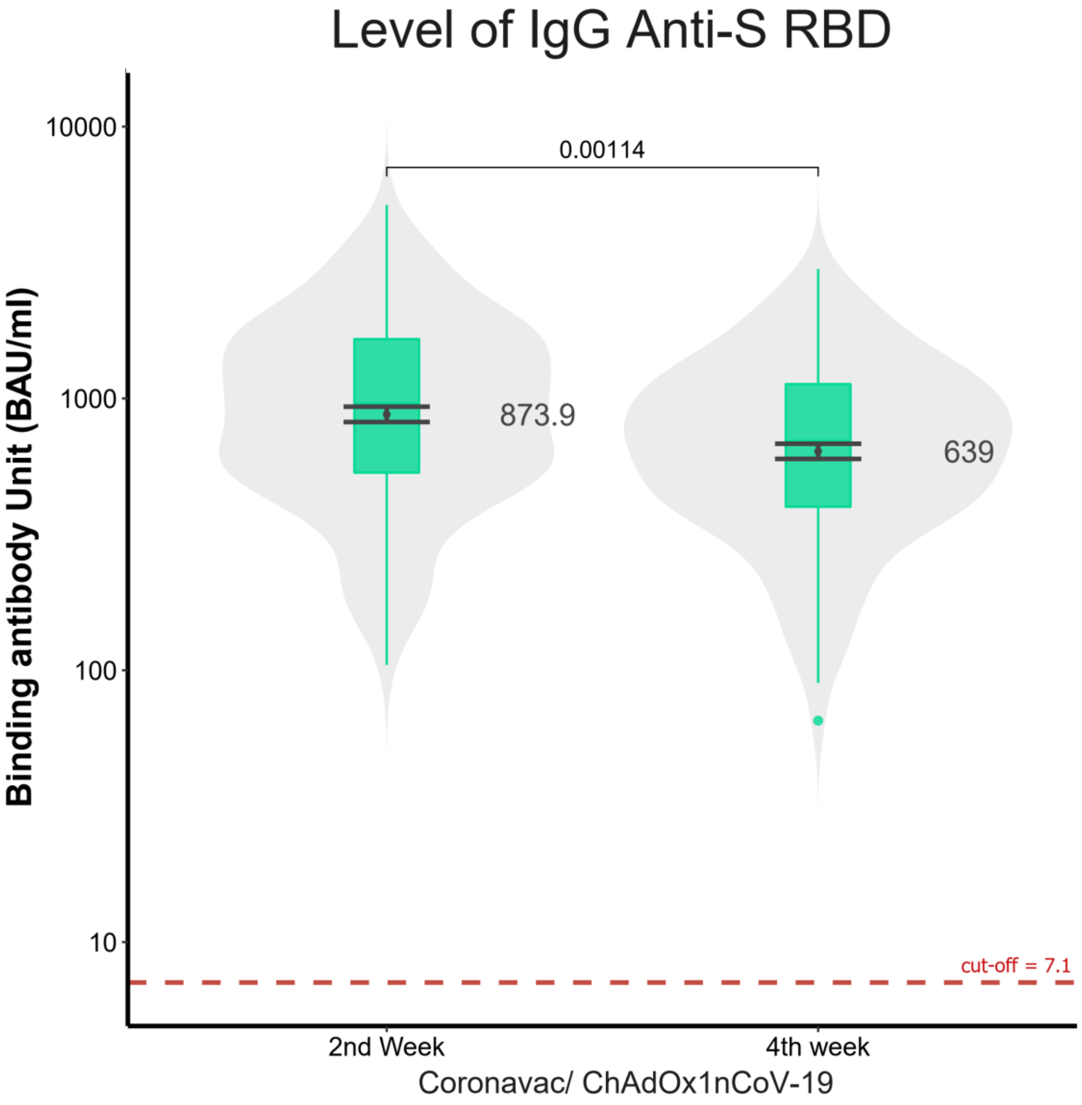


**Figure 4: Violin plots compare of level IgG anti-S RBD titer in I/V group at 2 wks and 4 wks. Boxplots show geomean and IQRs. level IgG anti-S RBD titer at 2 wks is significantly (p=0.00114) (873.9 BAU/mL, 95% CI 768.4-993.8) higher than 4 wks ( 639 BAU/mL, 95% CI 63-726). By using the non-parametric the Mann–Whitney U test.**

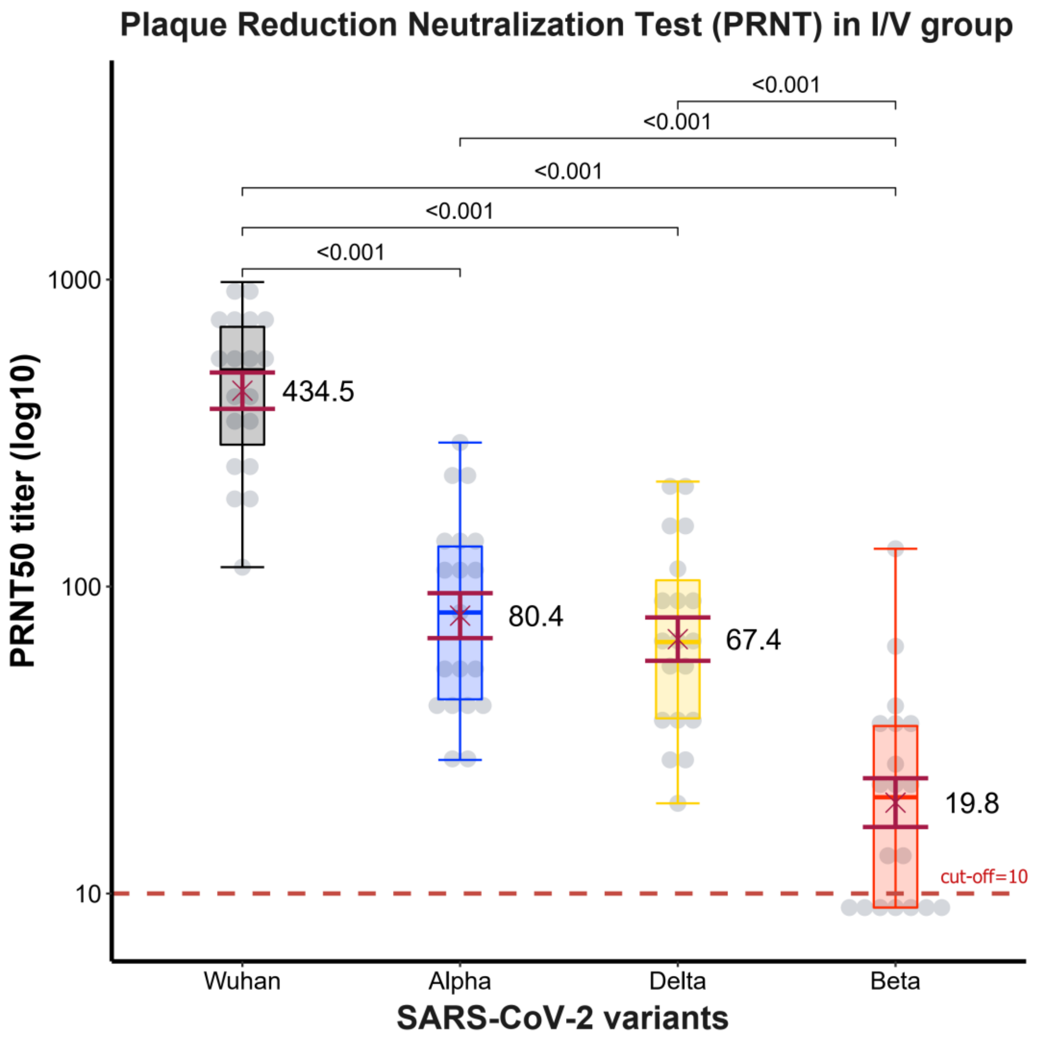


**Figure 5 : Boxplots indicate geomean $PRNT_{50}$ titer to virus variant and IQRs.serum collection at 2 wks after participants in the I/V group receive the vaccination. the GMT of $PRNT_{50}$ was the highest against Wuhan or wild type ( 434.5 BAU/mL, 95% CI 326-579) that significantly (p<0.001)greater than to Alpha variant (80.4 BAU/mL, 95% CI 56-115) , Delta variant( 67.4 BAU/mL, 95% CI 48-95) (p<0.001) , and Beta variant( 19.8 BAU/mL, 95% CI 14-30) (p<0.001).The horizontal dotted line indicates the positive detection (10 units). Use The Wilcoxon signed-rank test for assessment.**

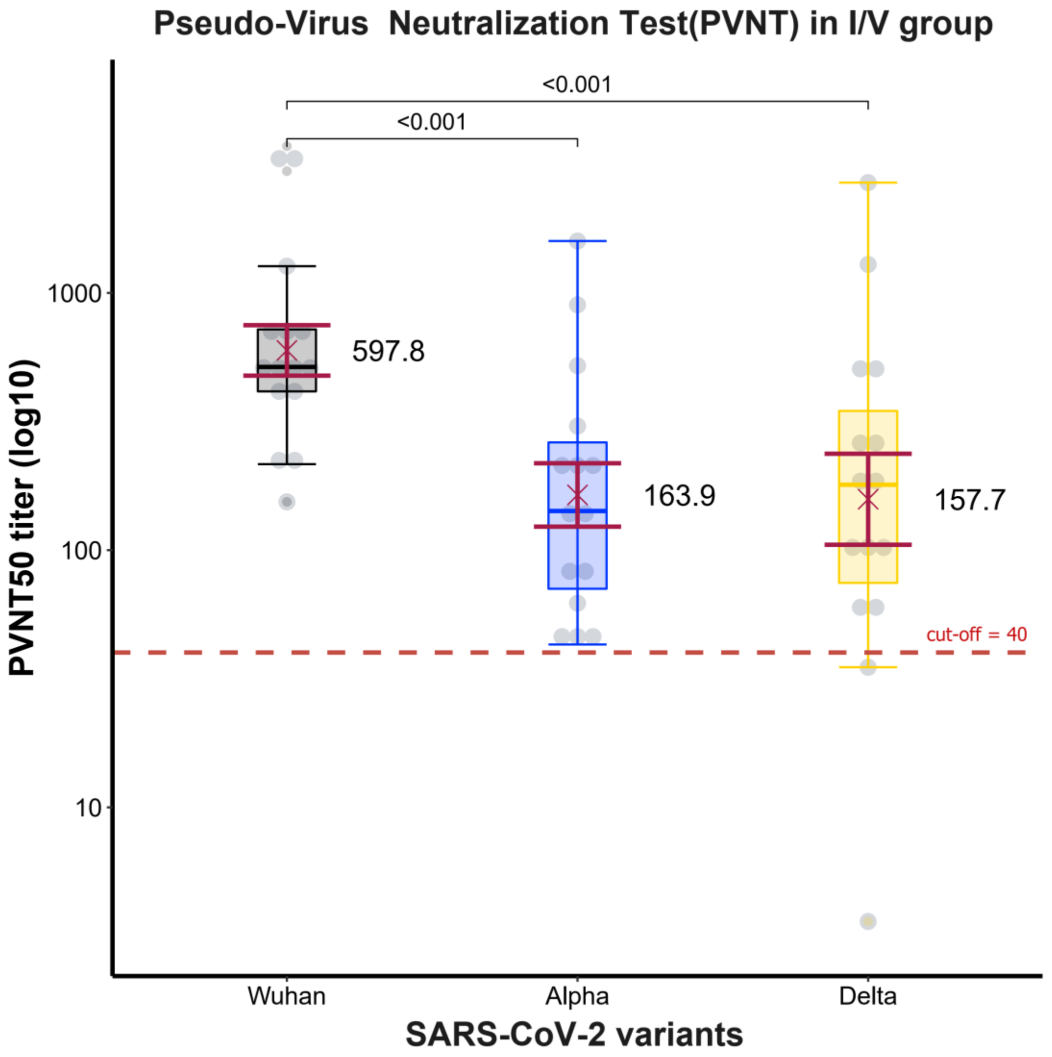


**Figure 6 : Boxplots indicate geomean $PVNT_{50}$ titer to virus variant and IQRs. Serum collection at 2 wks after participants in the I/V group receive the vaccination. the GMT of $PVNT_{50}$ was the highest against Wuhan or wild type (597.8 BAU/mL, 95% CI 326-579) that significantly ($p<0.001$)greater than to Alpha (163.9 BAU/mL, 95% CI 89-301), Delta variant( 157.7 BAU/mL, 95% CI 66-378) ($p<0.001$). The horizontal dotted line indicates the positive detection (40 units). Using the Wilcoxon signed-rank test for assessment.**

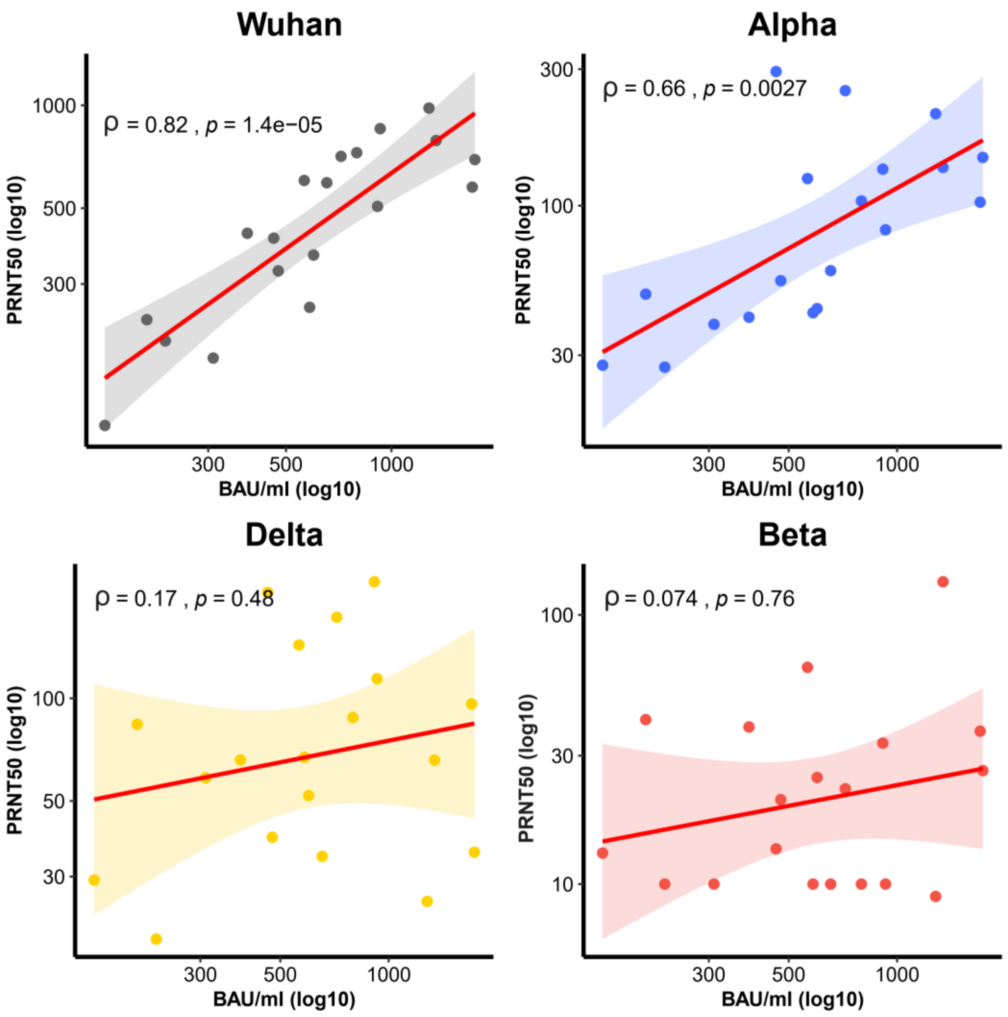


+

**Figure 7 : Correlation between $PRNT_{50}$ titer with Ab to anti-s RBD level by virus variants group**

**correlation (Solid line),95% confidence intervals( curved line) .Wild type (ρ =0.82,p<0.001), Alpha variant (ρ =0.66,p=0.0027), and Beta variant (ρ =0.074,p=0.76), Delta variant(ρ =0.17,p=0.48). The Spearman's rank correlation coefficient was used to assessment. Serum collection at 2 wks after participants in the I/V group receive a vaccination.**

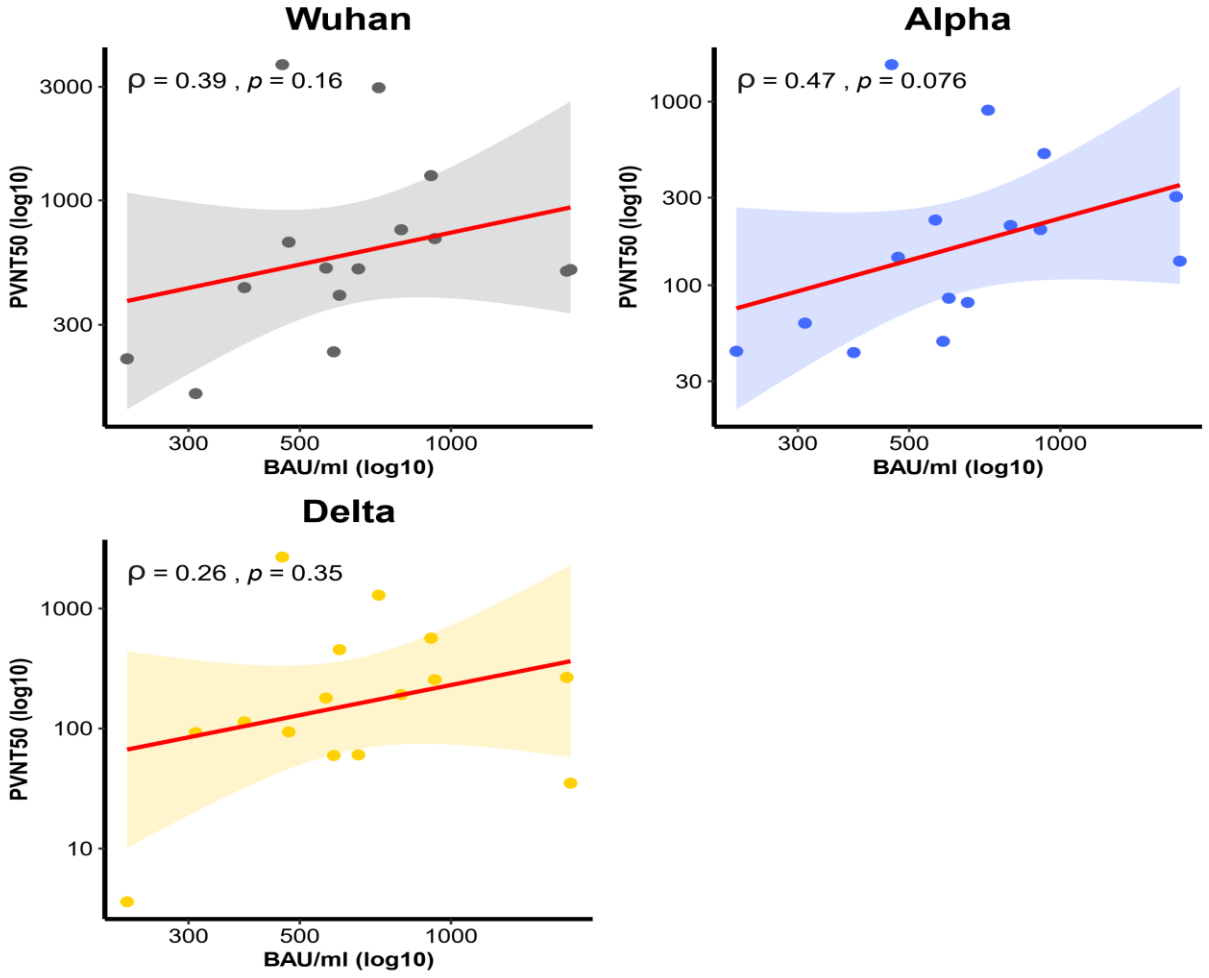


**Figure 8 : Correlation between $PVNT_{50}$ titer with Ab to anti-s RBD level by virus variants group (Wild type, Alpha variant, Delta variant) correlation (Solid line),95% confidence intervals(curved line)**

**Wild type (ρ =0.39, p=0.16), Alpha variant (ρ =0.47, p=0.076), Delta variant (ρ =0.26,p=0.35).**

**The Spearman's rank correlation coefficient was used to assessment. Serum collection at 2 wks after participants in the I/V group receive a vaccination.**

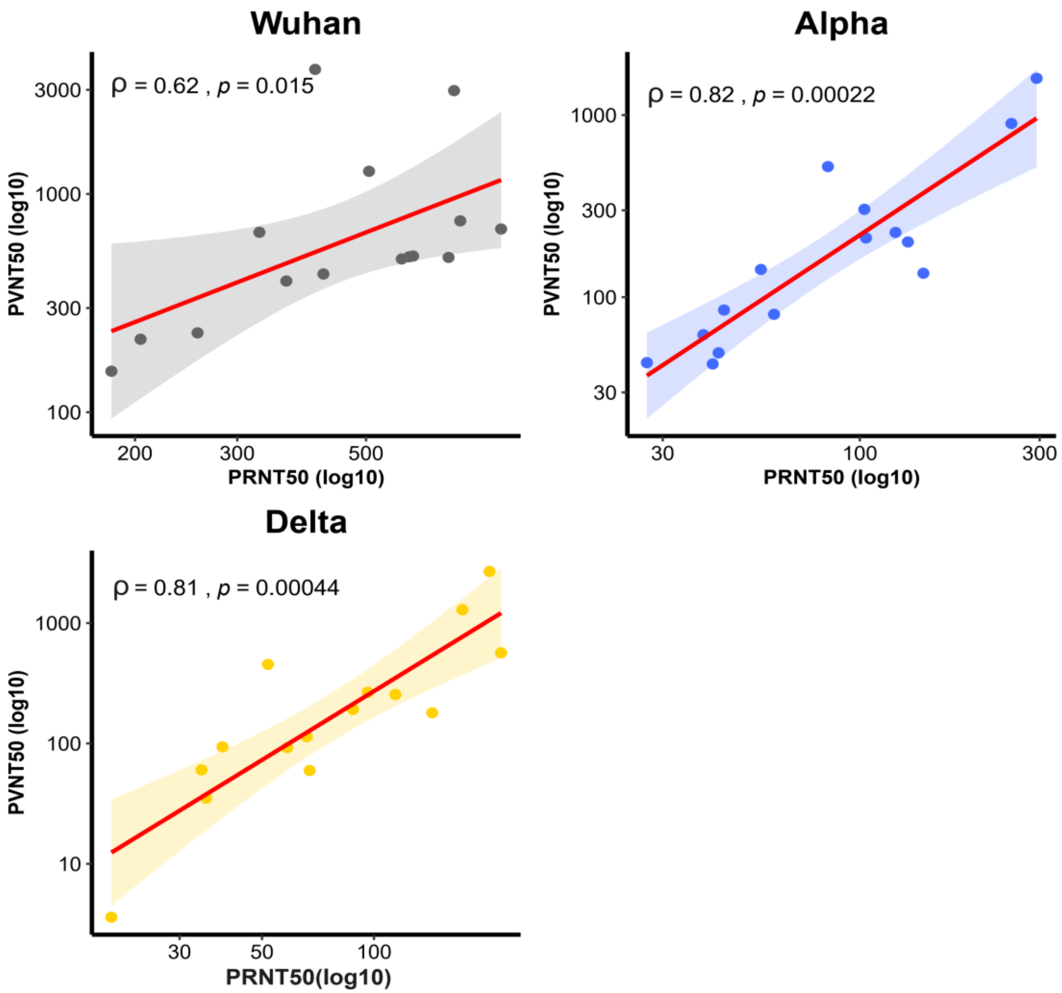


**Figure 9 : Correlation between $PVNT_{50}$ titer with $PRNT_{50}$ titer by virus variants group (Wild type, Alpha variant, Delta variant) correlation (Solid line),95% confidence intervals (curved line)**

**the correlation were Wild type (ρ =0.62, p=0.015), Alpha variant (ρ =0.82, p=0.00022), and Delta variant(ρ =0.81,p=0.00044).The Spearman's rank correlation coefficient was used to assessment. Serum collection at 2 wks after participants in the I/V group receive a vaccination.**
