## Supplementary table 1-9 for "Immunogenicity and adverse events of priming with inactivated whole SARS-CoV-2 vaccine (CoronaVac) followed by boosting the ChAdOx1 nCoV-19 vaccine"

TABLE 1: Demographic data of the study population 1
TABLE 2: Adverse event to first vaccine dose with coronavac in 30 participants from bangsue vaccination center 2
TABLE 3: Adverse event to first vaccine dose with coronavac in 125 participants from moph vaccination center 4
TABLE 4: Adverse event with in 2 wks following second dose with ChAdOx1 nCoV-19 5
TABLE 5: Show level of IgG anti-S RBD (bau/ml) at 4 wks 6
TABLE 6: Show level of IgG anti-S RBD (bau/ml) compare 2 wks with 4 wks 7
TABLE 7: Show geometric mean,geosd,median,IQRS,min – max, 95% ci of g.m. of $PRNT_{50}$ 8
TABLE 8: Show geometric mean,geosd,median,IQRS,min – max, 95% ci of g.m. of $PVNT_{50}$ 9
TABLE 9: Show percentage of participants with $PRNT_{50}$ titers and $PVNT_{50}$ titers 10

**Table 1: Demographic data of the study population**

| | GROUP 1A[1] Inactivated[3]/vector[4] | GROUP 1B[2] Inactivated/vector | Inactivated/ Inactivated | Vector/ vector | NI[5] |
|---|---|---|---|---|---|
| SEX | MOPH - VC | BVC | DMSC | ISDT | LAK SI |
| MALE | 61 | 15 | 6 | 7 | 80 |
| FEMALE | 64 | 15 | 26 | 40 | 89 |
| TOTAL | 125 | 30 | 32 | 47 | 169 |
| AGE GROUP | | | | | |
| 18-49 | 99 | 25 | 24 | 28 | 137 |
| MALE | 45 | 13 | 2 | 3 | 63 |
| FEMALE | 54 | 12 | 22 | 25 | 74 |
| 50-70 | 26 | 5 | 8 | 19 | 32 |
| MALE | 16 | 2 | 4 | 4 | 17 |
| FEMALE | 10 | 3 | 4 | 15 | 15 |
| MEDIAN AGE GROUP±SD | 40±8.8 | 40±9.24 | 45±8.8 | 53.5±14.6 | 38±11.7 |
| HISTORY COVID 19 INFECTION | 0 | 0 | 0 | 0 | 169 |

[1]125 participant at MOPH-VC , [2]30 participant at BVC,[3]ChAdOx1 nCoV-19, [4]CoronaVac**,** [5]participant from Laksi distric with history covid-19 infection **2021**

**Table 2: Adverse events to first vaccine dose with CoronaVac in 30 participants from Bangsue Vaccination Center**

| | Gender | Age range | Reason for switching of second vaccine to ChAdOx1 | Weeks between 1° and 2° vaccination, |
|---|---|---|---|---|
| 1. | Male | 36-40 | ISRR[1] -paresthesia numbness type grade2 | 3 wk |
| 2. | Male | 36-40 | urticaria grade 3 | 3 wk |
| 3. | Female | 26-30 | urticaria grade 3 | 3 wk |
| 4. | Female | 41-45 | maculopapular rash grade 1 | 3 wk |
| 5. | Female | 31-35 | ISRR -dizziness grade2 | 3 wk |
| 6. | Male | 26-30 | anaphylaxis grade 3 | 3 wk |
| 7. | Male | 26-30 | dizziness grade 1 | 3 wk |
| 8. | Male | 31-35 | chest wall pain grade 1 | 3 wk |
| 9. | Male | 36-40 | patient request | 3 wk |
| 10. | Female | 26-30 | dyspnea grade 2 | 3 wk |
| 11. | Male | 36-40 | anaphylaxis grade 3 | 3 wk |
| 12. | Male | 41-45 | angioedema/ diarrhea | 3 wk |
| 13. | Male | 31-35 | ISRR -paresthesia numbness type grade2 | 3 wk |
| 14. | Female | 51-55 | nausea vomiting/ malaise/ headache all grade 1 | 2 wks 5 days |
| 15. | Female | 41-45 | Injected site pain grade 1, angioedema grade 1 | 3 wks |
| 16. | Female | 31-35 | anaphylaxis grade 3 | 3 wks 2 days |

| | | | | |
|---|---|---|---|---|
| 17. | Male | 41-45 | patient request | 3 wks |
| 18. | Female | 61-65 | Injected site pain grade 1 | 3 wks |
| 19. | Female | 41-45 | ISRR - paresthesia numbness type grade2 | 3 wks 5 days |
| 20. | Female | 41-45 | maculopapular rash grade 1 | 3 wks |
| 21. | Male | 36-40 | central venous sinus thrombosis | 5 wks |
| 22. | Female | 46-50 | patient request | 3 wks |
| 23. | Male | 26-30 | Injected site pain grade 1, dyspnea grade 1, feeling feverish | 3 wks |
| 24. | Female | 46-50 | patient request | 2 wks 3 days |
| 25. | Female | 51-55 | ISRR blurred vision grade 2 | 6 wks |
| 26. | Female | 41-45 | anaphylaxis grade 3 | 5 wks |
| 27. | Male | 56-60 | ischemia cerebrovascular moderate symptom (Stroke) | 5 wks 6 days |
| 28. | Female | 26-30 | anaphylaxis grade 3 | 5 wks |
| 29. | Male | 51-55 | patient request | 3 wks 4 days |
| 30. | Male | 36-40 | hearing impairment grade 2 | 6 wks |
| | | | mean±sd | 3.5 **wks**±1.0 |

[1] "Immunization stress-related response" (ISRR) is a response to the stress some individuals may feel about getting an injection, and encompasses the spectrum of manifestations mentioned previously. The response to a stress encompasses a range of manifestations (symptoms and signs) that may include an acute stress response which includes a vasovagal reaction (fainting), hyperventilation or a dissociative neurological symptom reaction which includes non-epileptic seizures (formerly known as a conversion reaction).

[2]Grade refers to the severity of the AEs from Common Terminology Criteria for Adverse Events version 5.0 by the United States National Cancer Institute (NCI/NIH).

**Table 3: Adverse events to first vaccine dose with CoronaVac in 125 participants from MOPH Vaccination Center**

| | | Severity | | | | | |
|---|---|---|---|---|---|---|---|
| Adverse Events | N | Mild | | Moderate | | Severe | |
| | | N | % | N | % | N | % |
| feeling feverish | **125** | **8** | **6.40** | **0** | **0.00** | **0** | **0.00** |
| headache | **125** | **2** | **1.60** | **0** | **0.00** | **0** | **0.00** |
| injected site pain | **125** | **1** | **0.80** | **0** | **0.00** | **0** | **0.00** |
| dyspnea | **125** | **1** | **0.80** | **0** | **0.00** | **0** | **0.00** |
| nausea/vomitting | **125** | **1** | **0.80** | **0** | **0.00** | **0** | **0.00** |
| numbness | **125** | **0** | **0.00** | **0** | **0.00** | **0** | **0.00** |
| weakness | **125** | **1** | **0.80** | **0** | **0.00** | **0** | **0.00** |
| malaise/fatigue/hypersomnia | **125** | **5** | **4.00** | **0** | **0.00** | **0** | **0.00** |
| myalgia | **125** | **2** | **1.60** | **0** | **0.00** | **0** | **0.00** |
| petechiae | **125** | **0** | **0.00** | **0** | **0.00** | **0** | **0.00** |
| erythema | **125** | **1** | **0.80** | **0** | **0.00** | **0** | **0.00** |
| diarrhea | **125** | **2** | **1.60** | **0** | **0.00** | **0** | **0.00** |

**Table 4: Adverse events with in 2 wks following second dose with Chadox1 nCov-19 in 155 participants recieved CoronaVac/ChAdOx1 nCoV-19 regimen from MOPH-VC,BVC**

| Adverse Events | N | Severity | | | | | |
|---|---|---|---|---|---|---|---|
| | | Mild | | Moderate | | Severe | |
| | | N | % | N | % | N | % |
| feeling feverish | **155** | **103** | **66.45** | **1** | **0.65** | **0** | **0.00** |
| headache | **155** | **51** | 32.90 | **0** | 0.00 | **0** | 0.00 |
| injected site pain | **155** | **54** | **34.84** | **0** | **0.00** | **0** | **0.00** |
| dyspnea | **155** | **0** | 0.00 | **2** | 1.29 | **0** | 0.00 |
| nausea/vomitting | **155** | **13** | **8.39** | **0** | **0.00** | **0** | **0.00** |
| numbness | **155** | **1** | 0.65 | **0** | 0.00 | **0** | 0.00 |
| weakness | **155** | **0** | **0.00** | **0** | **0.00** | **0** | **0.00** |
| malaise/fatigue/hypersomnia | **155** | **30** | 19.35 | **0** | 0.00 | **0** | 0.00 |
| myalgia | **155** | **40** | **25.81** | **0** | **0.00** | **0** | **0.00** |
| petechiae | **155** | **0** | 0.00 | **0** | 0.00 | **0** | 0.00 |
| erythema | **155** | **3** | **1.94** | **0** | **0.00** | **0** | **0.00** |
| diarrhea | **155** | **4** | **2.58** | **0** | **0.00** | **0** | **0.00** |
| poor appetize | **155** | **1** | 0.65 | **0** | 0.00 | **0** | 0.00 |
| chill | **155** | **1** | 0.65 | **0** | 0.00 | **0** | 0.00 |
| urticaria | **155** | **1** | 0.65 | **0** | 0.00 | **1** | 0.65 |

**Table 5: Show Level of IgG Anti-S RBD (BAU/ml) in mean, Geometric mean,GeoSD,median ,IQRs Min-max,95% CI of G.M. at 4 wks.**

| | Level of IgG Anti-S RBD (BAU/ml) | | | |
|---|---|---|---|---|
| | **Natural infection** | **Coronavac/Coronavac** | **ChAdOx1 nCoV-19 / ChAdOx1 nCoV-19** | **Coronavac/ ChAdOx1 nCoV-19** |
| **n** | 169 | 32 | 47 | 137 |
| **mean** | 449.9 | 160 | 308.5 | 819.5 |
| **geoMean** | 177.3 | 108.2 | 211.1 | 639 |
| **geoSD** | 4.3 | 2.6 | 2.5 | 2.1 |
| **median** | 185.8 | 117.1 | 214.4 | 690.4 |
| **Q1-Q3** | 59.9-471.8 | 55.9-205.5 | 128.7-354.5 | 399.3-1129.4 |
| **min-max** | 8.8-3320.7 | 10.9-738.7 | 9.5-1787.5 | 65.3-2996.2 |
| **95% CI of G.M.** | 42-221 | 77-152 | 162-249 | 63-726 |

**Table 6: Show Level of IgG Anti-S RBD (BAU/ml) in mean, Geometric mean,GeoSD,median ,IQRs Min-max,95% CI of G.M. at 2 wks**

| | LEVEL OF IgG ANTI-S RBD (BAU/ML) | |
|---|---|---|
| | 2nd Week | 4th week |
| **N** | 149 | 137 |
| **MEAN** | 1164.0 | 819.5 |
| **GEOMEAN** | 873.9 | 639.0 |
| **GEOSD** | 2.2 | 2.1 |
| **MEDIAN** | 947.9 | 690.4 |
| **Q1-Q3** | 533.3-1653.2 | 399.3-1129.4 |
| **MIN-MAX** | 104.6-5150.5 | 65.3-2996.2 |
| **95%CI OF G.M.** | 768.4-993.8 | 63-726 |

**Table 7: Show Geometric mean,GeoSD,median,IQRs,min – max, 95% CI of G.M. of $PRNT_{50}$ from 19 participants in CoronaVac/ChAdOx1 nCoV-19 regimen at 2 wks.**

| $PRNT_{50}$ titers | Wild type | Alpha | Delta | Beta |
|---|---|---|---|---|
| n | 19 | 19 | 19 | 19 |
| mean | 502.7 | 103.7 | 85 | 27.9 |
| geoMean | 434.5 | 80.4 | 67.4 | 19.8 |
| geoSD | 1.8 | 2.1 | 2 | 2.2 |
| median | 506.09 | 82.31 | 65.97 | 20.59 |
| Q1-Q3 | 292.0-701.5 | 42.9-135.1 | 37.3-105.2 | 9.0-35.2 |
| min-max | 115.6-981.6 | 27.2-294.4 | 19.7-219.8 | 9.0-132.8 |
| 95% CI of G.M. | 326-579 | 56-115 | 48-95 | 14-30 |

**Table 8: Show Geometric mean,GeoSD,median,IQRs,min – max, 95% CI of G.M. of PVNT$_{50}$ from 15 participants in CoronaVac/ChAdOx1 nCoV-19 regimen at 2 wks.**

| PVNT$_{50}$ titers | Wild type | Alpha | Delta |
|---|---|---|---|
| **n** | 15 | 15 | 15 |
| **mean** | 306.4 | 423.1 | 903.9 |
| **geoMean** | 163.9 | 157.7 | 597.8 |
| **geoSD** | 3 | 4.8 | 2.4 |
| **median** | 142 | 179.6 | 515.4 |
| **Q1-Q3** | 71.4-265.2 | 76.3-360.2 | 414.5-721.9 |
| **min-max** | 43.0-1591.4 | 3.6-2684.3 | 154.1-3720.7 |
| **95% CI of G.M.** | 368-970 | 89-301 | 66-378 |

**Table 9: Show [1]Percentage of participants with $PRNT_{50}$ titers ≥10 (NAb positivity cut-off) against wild type,Alpha, Beta and Delta strains .**

**[2]Percentage of participants with $PVNT_{50}$ titers ≥40 (NAb positivity cut-off) against wild type,Alpha and Delta strains.**

| | WILD TYPE | ALPHA | DELTA | BETA |
|---|---|---|---|---|
| **[1]$PRNT_{50}$ TITER >10** | 19/19(100%) | 19/19(100%) | 19/19(100%) | 12/19(63%) |
| **[2]$PVNT_{50}$ TITER >40** | 15/15(100%) | 15/15(100%) | 13/15(86.67%) | n/a |
